# Clinical Results with a B Cell Activating Anti-CD73 Antibody for the Immunotherapy of COVID-19

**DOI:** 10.1101/2021.09.13.21263406

**Authors:** Richard A. Miller, Pramod Guru, Philippe Bauer, Jorge Robles, Christian Tomaszewski, J. Scott Overcash, Michael Waters, Miriam Cameron, Julián Olalla Sierra, Haider Mashhedi, Mehrdad Mobasher, James Janc, Jenny A. Rudnick, Shenshen Hu, William B. Jones, Long Kwei, Suresh Mahabhashyam, Stephen B. Willingham, Gerard Criner

**Affiliations:** Corvus Pharmaceuticals, Burlingame, CA, USA; Mayo Clinic, Jacksonville, FL, USA; Mayo Clinic, Rochester, MN, USA; El Centro Regional Medical Center, El Centro, CA, USA; University of California, San Diego, CA, USA; Sharp Grossmont Hospital, La Mesa, CA, USA; Sharp Chula Vista Medical Center, Chula Vista, CA, USA; Holy Cross Hospital, Silver Spring, MD, USA; Hospital Costa del Sol, Marbella, Spain; Temple University Hospital, Philadelphia, PA, USA

**Keywords:** CD73, COVID-19, SARS-CoV-2, RBD, vaccine, antibody, B cells, T cells

## Abstract

Robust polyclonal humoral immune responses have the potential to generate a diverse set of antibodies to neutralize and eliminate viruses such as SARS-CoV-2 and protect against transmission, re-infection and the evolution of variants that evade immunity. CD73 is present on subsets of human B and T cells where it plays a role in lymphocyte activation and migration. CD73 also functions as an ectoenzyme that converts AMP into immunosuppressive adenosine. We have developed a humanized anti-CD73 antibody, mupadolimab (CPI-006), that blocks CD73 enzymatic activity and activates CD73^POS^ B cells, thereby inducing differentiation into plasmablasts, immunoglobulin class switching, and antibody secretion independent of the adenosine modulatory activity. These effects suggest mupadolimab may enhance the magnitude, diversity, and duration of anti-viral responses in patients with COVID-19. This hypothesis was tested in a dose escalation phase 1 trial in 29 hospitalized patients with COVID-19. Single doses of 0.3 mg/kg – 5 mg/kg mupadolimab were well tolerated with no drug related adverse events. Doses greater than 0.3 mg/kg resulted in rapid generation of IgG and IgM to SARS-CoV-2 significantly above titers measured in convalescent controls, with elevated IgG titers sustained for more than 6 months beyond presentation of symptoms. Based on these findings, a randomized double-blind, placebo-controlled Phase 3 study in hospitalized patients was initiated. The primary endpoint was proportion of patients alive and free from respiratory failure within 28 days. This trial was discontinued early during the period of waning COVID-19 incidence after enrolling 40 patients. Although underpowered, results from this trial suggest improvement in the primary and key secondary endpoints in patients treated with single doses of 2 mg/kg and 1 mg/kg compared to placebo. The presumed mechanism of action, stimulation of B cells, may represent a novel approach to immunotherapy of COVID-19 and other viral infections.

## INTRODUCTION

COVID-19 is a viral disease caused by the severe acute respiratory syndrome coronavirus 2 (SARS-CoV-2). Despite the introduction of vaccines and passive monoclonal anti-viral antibodies, there remains an urgent need for therapies that can improve survival, clinical outcomes, and reduce the requirements for intensive supportive care and prolonged hospitalization.[1-3] Recent emergence of viral variants and deterioration of immunity with time following vaccination have highlighted these needs. In addition, immunocompromised individuals have shown vulnerability to infection despite vaccination.[4] There is also evidence that chronic or persistent infection with SARS-CoV-2 may lead to variants with the potential to escape immunity. Anti-inflammatory drugs such as dexamethasone, tocilizumab, baricitinib and tofacitinib have shown only slight clinical benefit in selected subgroups of hospitalized patients.[5-8] The inadequate results to date and continued surge in viral spread and disease, demonstrate the urgent need for new approaches to prevent, control or eradicate viral infection, especially those infections resulting in persistent or serious disease.

CD73 is expressed on subsets of human CD4^POS^ and CD8^POS^ T cells, germinal center follicular dendritic cells, and both naïve and class switched memory B cells where it is thought to play a role in lymphocyte trafficking and cellular activation.[9-14] CD73 also functions as an ectonucleotidase that hydrolyzes extracellular adenosine monophosphate (AMP) into immunosuppressive adenosine.[15]

Mupadolimab is an IgG1κ humanized FcγR binding-deficient anti-CD73 monoclonal antibody (mAb) that activates CD73^POS^ B cells.[16, 17] In vitro, mupadolimab induces the increased expression of markers associated with B cell maturation and antigen presentation, morphologic transformation to plasmablasts, and increased secretion of IgM and IgG. The unique immunologic properties of mupadolimab provided the rationale to examine its use as an immunotherapy for COVID-19 with the aim of boosting anti-viral immune responses and improving clinical outcome. We conducted an open label phase 1 trial with mupadolimab in hospitalized patients with COVID-19. Findings in this study led to the initiation of a randomized double-blind, placebo-controlled phase 3 study that was terminated early after enrollment of 40 patients due to the waning incidence of COVID-19 in the U.S. in mid 2021. The clinical results of these trials are reported here. The findings suggest that immune-stimulation with mupadolimab offers a novel approach to treating COVID-19, and potentially other infectious diseases, which relies on the stimulation of B cells in the presence of antigens from an infectious agent to enhance immunity.

## RESULTS

### Mupadolimab Elicits Antigen Specific Immunity in an Animal Model

We utilized an animal model to determine if mupadolimab could enhance antigen-specific immune responses to SARS-CoV-2. Mupadolimab does not bind to mouse CD73, so NSG-SGM3 mice were used. Though these mice have no mouse B, T, or NK cells, they have been reconstituted with a human immune system and do have human B cells that can be activated with mupadolimab.[18, 19] NSG-SGM3 mice were immunized with purified SARS-CoV-2 trimeric spike protein (TS) in incomplete adjuvant, along with mupadolimab or a human IgG1 isotype control. Mice vaccinated with the TS plus mupadolimab made antigen specific human anti-TS antibodies, while mice receiving TS plus isotype control did not mount a response (Figure 1A). These antibody responses were antigen specific, as mice treated with mupadolimab made antibodies to the immunizing TS protein; but not to the control nucleocapsid SARS-CoV-2 viral protein (Figure 1B). These results demonstrate that mupadolimab stimulates antigen specific humoral immunity and would be expected to potentiate antibody responses to SARS-CoV-2 in patients with COVID-19.

**Fig 1.**
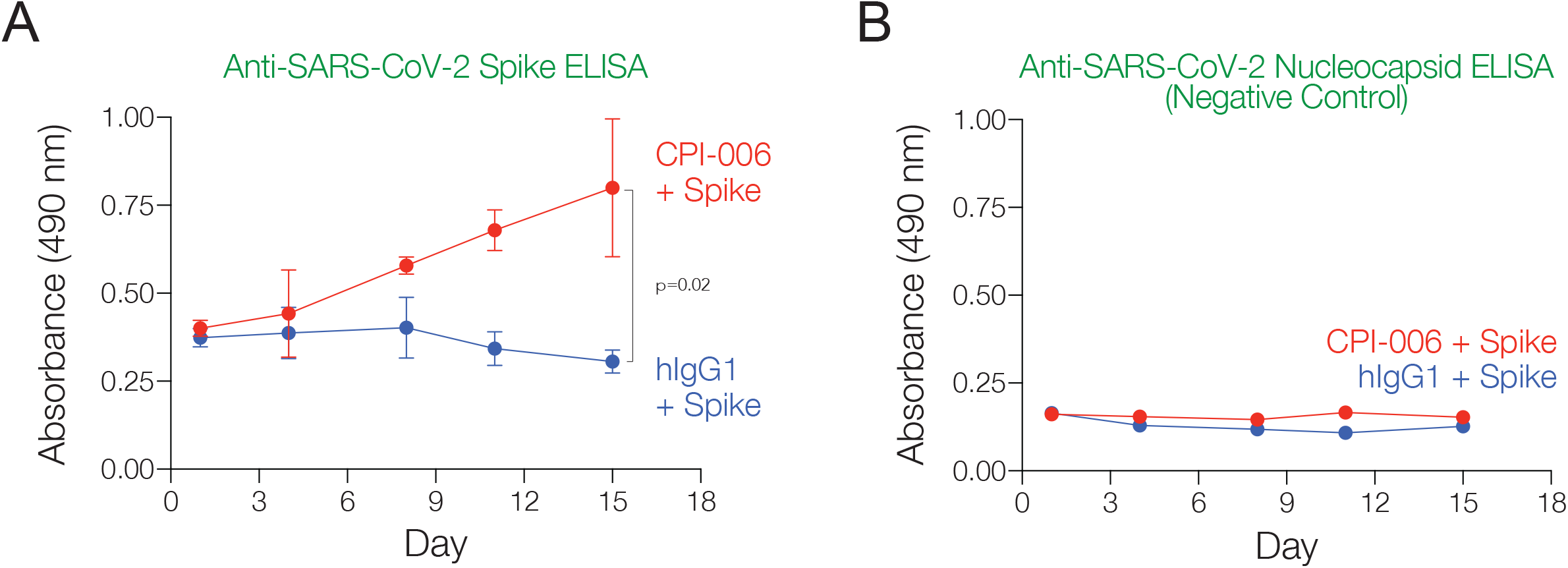
Mupadolimab Elicits Antigen Specific Immunity in a Preclinical Animal Model. NSG-SGM3 mice were immunized with an emulsion of 50 μg full length spike protein from SARS-CoV-2 plus Freund’s incomplete adjuvant (subcutaneously on both the left and right flank (25 μg/side). Mice were then randomized into two treatment groups: half the animals were dosed daily (i.p.) with 10 mg/kg mupadolimab, and half the animals were dosed daily with 10 mg/kg hIgG1. Animals were cheek bled on days 1 (pre-treatment), 4, 8, 11, and 15 to assess anti-Spike (A) or anti-nucleocapsid (B, as a negative control) antibody production over time by ELISA.

### Immunotherapy of COVID-19

#### Phase 1 open label trial

We conducted a Phase 1 open label, single dose-escalation trial evaluating the safety, immunologic effects and clinical outcomes in COVID-19 patients hospitalized with mild-moderate disease (NCT04464395). SARS-CoV-2 infection was confirmed by RT-qPCR (reverse transcriptase quantitative polymerase chain reaction) testing of nasal swabs and eligible patients had blood oxygen saturation of at least 92% on ≤5L/min supplemental oxygen. Cohorts of patients received doses of 0.3 mg/kg, 1.0 mg/kg, 2.0 mg/kg, 3.0 mg/kg or 5.0 mg/kg administered by intravenous infusion given over 5-10 minutes. Patients in the 2 mg/kg cohort were included after enrollment of the other cohorts in order to obtain additional clinical data related to dosing. Patients could receive standard care for COVID-19, including remdesivir and steroids. No patient received passive monoclonal antibody therapy and two patients received convalescent plasma. Safety and other disease assessments along with PBMC (peripheral blood mononuclear cell) and serum collection were conducted daily while patients were hospitalized and then at 7, 14, and 28 days, and at 2, 3, and 6 months after receiving mupadolimab. Clinical assessments and endpoints included time to hospital discharge and proportion of patients progressing to respiratory failure.

Table 1 shows the patient characteristics of 29 patients treated on the study. These patients all had unfavorable comorbidities or were from high risk populations. The median time post onset of symptoms (POS) to mupadolimab administration was 8 days (range 1-21). No drug related adverse events of any grade or changes in quantitative serum immunoglobulins were observed. All patients recovered with improvement of inflammatory markers and symptoms and were discharged at a median of 3.0 days after hospitalization (Table 1). No patients progressed to requiring invasive or non-invasive mechanical ventilation.

**Table 1:** Baseline characteristics of COVID-19 patients treated in the Phase 1. CAD - coronary artery disease, CKD - chronic kidney disease, COPD - chronic obstructive pulmonary disease, DM - diabetes, HTN - hypertension

| Dose (mg/kg) | # of Patients Enrolled | Median Age (Range) | Onset of Symptoms Median (Range) | Comorbidities | Median BMI (Range) | Median Time to Discharge (Range) |
| --- | --- | --- | --- | --- | --- | --- |
| 0.3 | 5 | 48 (28-72) | 4 (1-8) | DM, CAD, HTN, asthma, cancer | 30.3 (24.6-33.7) | 3 (2-4) |
| 1.0 | 11 | 67 (37-80) | 7 (3->21) | DM, CAD, HTN, COPD, hypothyroidism | 32.1 (16.5-40.1) | 4 (2-13) |
| 2.0 | 3 | 52 (47-85) | 5 (2-6) | DM, HTN, CAD, asthma | 36.7 (23.8-41.8) | 3 (2-4) |
| 3.0 | 5 | 53 (26-76) | 5 (1-9) | DM, HTN, asthma, cancer | 30.7 (26.5-33.9) | 4 (2-23) |
| 5.0 | 5 | 56 (23-68) | 5 (4-8) | DM, HTN, CKD, cancer | 33.3 (23.3-47.5) | 4 (3-8) |
| OVERALL | 29 | 61 (23-85) | 5 (1->21) |  | 32.1 (16.5-47.5) | 3 (2-23) |

#### Immune Responses in Mupadolimab Treated COVID-19 Patients

IgG antibody titers against the SARS-CoV-2 TS and/or RBD (receptor binding domain) were substantially increased in all evaluable patients 28 days after a single infusion of low doses of mupadolimab (Figure 2A-B). Lower antibody titers were seen in the cohort receiving 0.3 mg/kg, consistent with plasma concentrations of mupadolimab below that needed for maximum B cell activation. Serum concentrations of mupadolimab achieved levels exceeding 1 μg/ml for greater than 24 hours for doses of 1 mg/kg and higher; concentrations known to activate B cells in vitro.[17] IgG titers were sustained without decrement up to 6 months after POS (Figure 2C). Antibody levels reached higher titers compared to convalescent sera from recovered patients (convalescent sera collected at 4-6 weeks POS, no comorbidities, and median age of 40) (Figure 2A-B, 2D-E). Similar results in the magnitude and duration of anti-SARS-Cov-2 IgM titers (Figure 2D-E) were also observed with decreasing titers seen beyond 84 days. No correlation between time after POS and pre-treatment serum antibody levels was observed as all patients had low pre-treatment titers despite relatively long durations of COVID-19 related symptoms. Mupadolimab did not induce non-specific polyclonal antibody responses. No increase in serum antibodies to common viral antigens was observed, nor were autoantibodies to type 1 IFNs, previously associated with severe COVID-19, increased following mupadolimab treatment (data not shown).[20] Immunophenotyping of PBMCs at baseline and 14, 28, and 56-days after treatment provided preliminary evidence that mupadolimab increased the frequency of memory B cells (Figure 2F). No increase in B cells with a memory phenotype were observed in the lowest dose group (0.3 mg/kg).

**Fig 2.**
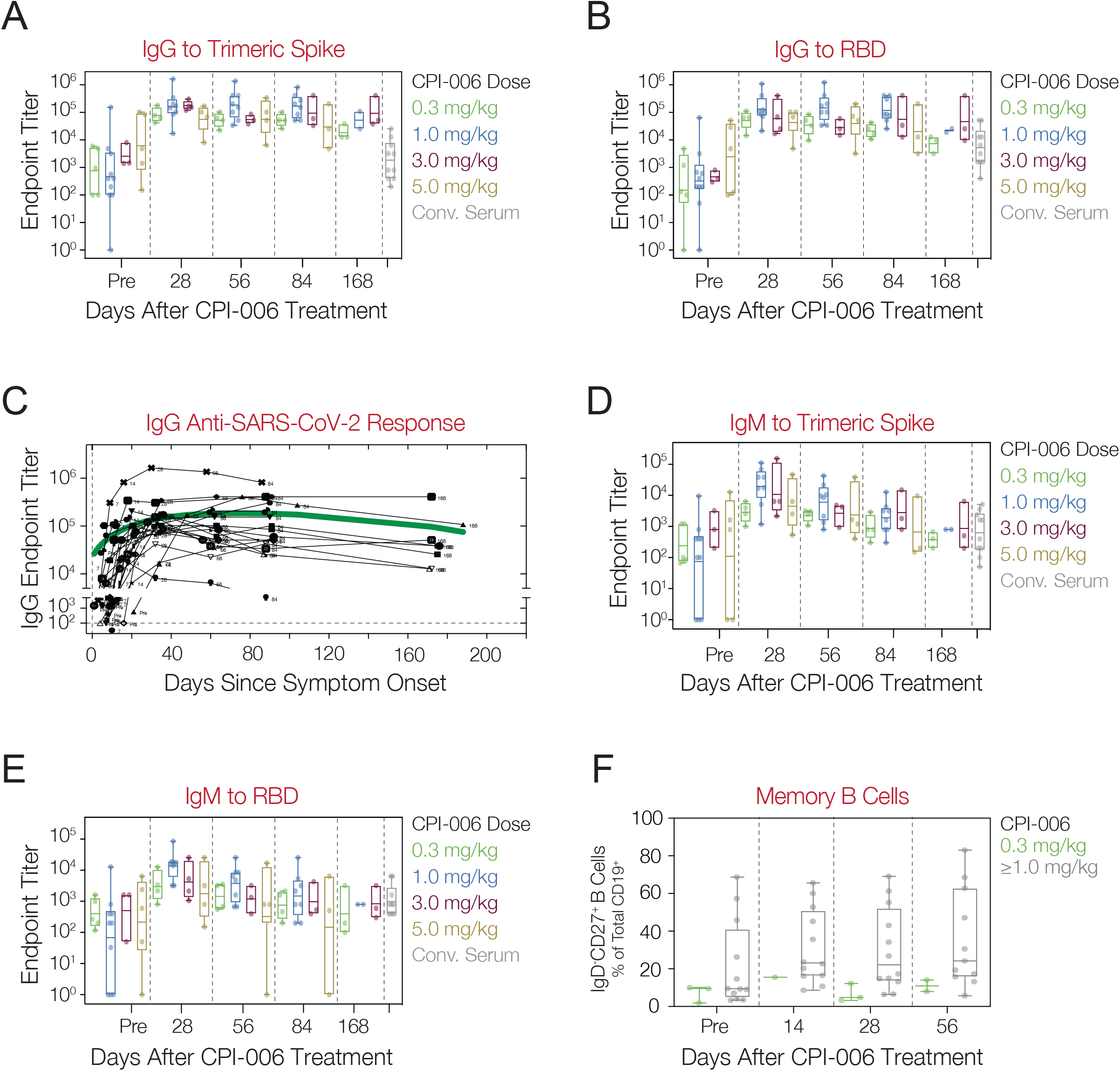
Anti-SARS-CoV-2 antibody and cellular responses in COVID-19 patients treated with mupadolimab. A, B) Patients received a 0.3, 1.0, 3.0 or 5.0 mg/kg single dose of mupadolimab and endpoint IgG titers to TS (A) and RBD (B) were measured at pre-treatment and at day 28, 56, 84, and 168. C) Longitudinal analyses of IgG against SARS-CoV-2 in mupadolimab treated subject. The green line is a smoothing spline fit. Each black line represents an individual patient. D, E) Patients received a 0.3, 1.0, 3.0 or 5.0 mg/kg single dose of mupadolimab and endpoint IgM titers to TS (D) and RBD (E) were measured at pre-treatment and at day 28, 56, 84, and 168. F) Frequency of circulating memory B cells (CD19^POS^IgD^NEG^CD27^POS^) within CD19^POS^ gate at baseline and after treatment in patients treated with 0.3 mg/kg mupadolimab compared to ≥1.0 mg/kg. Data are shown as box and whisker plot with geometric mean and interquartile range. Each dot represents a patient. Also shown are titers from convalescent patient serum obtained 4-6 weeks after POS.

We next evaluated if the anti-SARS-CoV-2 antibody responses following mupadolimab treatment correlated with increased neutralization activity in SARS-CoV-2 pseudotyped lentiviral infectivity assays. Prolonged and elevated neutralizing titers were observed in patients following mupadolimab treatment, with ID_50_ values up to 24,000 that persisted more than 56 days following POS (Figure 3A). Neutralizing antibody titers in patients treated with mupadolimab exceed titers reported for other hospitalized COVID-19 patients.[21, 22] We tested serum from 12 patients in the study enrolled prior to reported emergence of variants (Figure 3B). Day 28 serum from these mupadolimab treated patients effectively neutralized the B.1.1.7 variant. Nine of the 12 patients had neutralizing titers above 1:50 and 7 of the 12 patients had neutralizing titers of > 1:100 against the B.1.351 variant harboring both the E484K and N501Y mutations in the RBD that are reported to increase transmission and confer resistance to several neutralizing monoclonal antibodies in clinical development (Figure 3B).[23-25] These results suggest mupadolimab treatment can elicit a robust and durable polyclonal neutralizing antibody response in COVID-19 patients that is capable of providing cross-protection against emerging SARS-CoV-2 variants.

**Fig 3.**
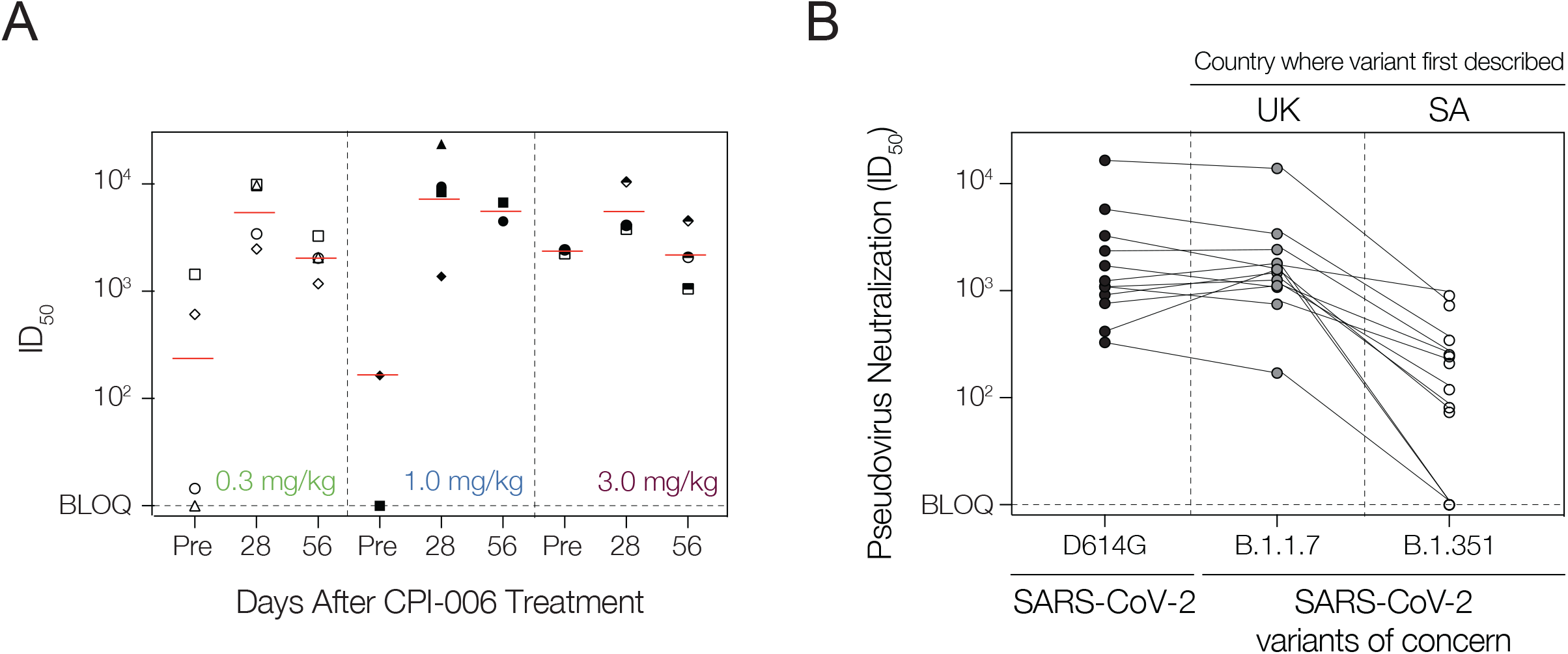
Sera from mupadolimab treated subjects neutralize wild type SARS-CoV-2 and spike variants. A) Titers required to achieve 50% neutralization (ID_50_) of infectivity with wild type SARS-CoV-2 pseudovirus. B) Neutralization (ID_50_) of infectivity with spike variant pseudovirus. Each symbol represents an individual patient. UK - United Kingdom, SA - South Africa.

### Randomized Double-Blind Placebo-Controlled Trial

#### Clinical design and endpoints

A Phase 3, randomized, 3-arm, placebo-controlled, double-blind, multicenter, stratified study of mupadolimab plus standard of care (SOC) versus placebo plus SOC in mild to moderately symptomatic hospitalized patients with COVID-19 was initiated. A planned 1000 participants would be randomized at a 1:1:1 ratio to the 3 treatment arms and stratified by the following factors: age, region of the world and comorbidities. The three treatment arms received intravenous mupadolimab 2 mg/kg, 1 mg/kg or placebo delivered by 5-10-minute infusion. COVID-19 disease was confirmed by RT-qPCR or antigen testing and symptoms compatible for SARS-CoV-2. Patients with signs of acute respiratory distress syndrome (ARDS) or respiratory failure necessitating mechanical ventilation at the time of screening (and randomization) or anticipated impending need for mechanical ventilation were excluded. The primary endpoint was to compare the proportion of participants alive and respiratory failure free during the 28 days after dosing with mupadolimab plus SOC versus placebo plus SOC. An 8-point ordinal disease assessment score was used to characterize patient illness as follows:

1. Not hospitalized, no limitations on activities
2. Not hospitalized, limitation on activities and/or requiring home oxygen
3. Hospitalized, not requiring supplemental oxygen - no longer requiring ongoing medical care
4. Hospitalized, not requiring supplemental oxygen - requiring ongoing medical care (COVID-19 related or otherwise)
5. Hospitalized, requiring supplemental oxygen
6. Hospitalized, on non-invasive ventilation or high flow oxygen devices
7. Hospitalized, on invasive mechanical ventilation or extracorporeal membrane oxygenation
8. Death

Only patients with score 4, 5, or 6 on the 8-point scale were admitted to the study.

Secondary endpoints included the time to clinical improvement, time to sustained recovery and time to discharge from the hospital during the 28 days after dosing. Day of recovery was defined as the first day on which the participant satisfied 1 of the following 3 categories from the 8-point ordinal scale 1) Not hospitalized, no limitations on activities.; 2) Not hospitalized, limitation on activities and/or requiring home oxygen; 3) Hospitalized, not requiring supplemental oxygen - no longer requires ongoing medical care. Time to clinical improvement required ≥ 2 points improvement in the 8-point ordinal scale during the 28 days after dosing. Sustained recovery required patients to be free of relapse or worsening after improvement.

The statistical design was based on approximately 330 participants per treatment arm where there would be an approximately 80% power to show a statistically significant superiority of 2 mg/kg dose over placebo in the proportion of participants alive and free from respiratory deterioration during the 28 days after dosing at a 1-sided alpha level of 0.0125 when true proportion of 2 mg/kg dose is 92% and placebo is 84%. The same sample size and power statement was used for the comparison between 1 mg/kg and placebo.

In addition to clinical endpoints, serum samples were collected at various times and were tested for antibody response to SARS-CoV-2 including several variants.

Enrollment began in February 2021. In July 2021, after enrolling 40 patients, it was decided to discontinue the phase 3 trial due to slower than expected enrollment and waning of the COVID-19 incidence in the U.S. before the 4th wave associated with the delta variant occurred. Given the importance of the pandemic and need for more effective therapies, we analyzed the available data recognizing the sample size was underpowered to establish statistical significance. The results in these 40 patients are reported below. Data from the three cohorts remained blinded until this analysis.

#### Clinical Results

Forty patients were randomized to receive standard of care plus either single dose: Mupadolimab 1 mg/kg (N=14), 2 mg/kg (N=15) or placebo (N=11) at 8 sites (7 U.S.). The clinical characteristics are shown in Table 2. Generally, the treatment arms were balanced. Overall, the median age was 55 years and 42% were female. Presenting symptoms were typical for patients with COVID-19. Patients in the 2 mg/kg cohort had more severe disease on admission with 60% having a score of 6 points on the 8-point ordinal scale vs. 7% in the 1 mg/kg and 27% in the placebo cohorts.

**Table 2:** Baseline characteristics of enrolled COVID-19 patients treated in the Phase 1. Baseline patient characteristics are shown. Comorbidities include: diabetes, hypertension, asthma, obesity, cancer, etc. BMI - body mass index

| Baseline characteristics | 2 mg/kg + SOC<br>(N=15) | 1 mg/kg + SOC<br>(N=14) | Placebo + SOC<br>(N=11) | Total<br>(N=40) |
| --- | --- | --- | --- | --- |
| **Age (yrs.) – Median (range)** | 56.0 (28, 69) | 53.0 (25, 67) | 53.0 (21, 63) | 55.0 (21, 69) |
| **Gender (Female) – n (%)** | 6 (40.0) | 3 (21.4) | 8 (72.7) | 17 (42.5) |
| **Race – n (%)** |  |  |  |  |
| White | 12 (80.0) | 11 (78.6) | 8 (72.7) | 31 (77.5) |
| African American | 2 (13.3) | 2 (14.3) | 2 (18.2) | 6 (15.0) |
| Asian | 0 | 1 (7.1) | 1 (9.1) | 2 (5.0) |
| Hispanic or Latino | 6 (40.0) | 3 (21.4) | 3 (27.3) | 12 (30.0) |
| **BMI – Median (range)** | 33.10 (19.5, 61.7) | 30.75 (20.4, 45.6) | 38.90 (23.7, 47.7) | 33.15 (19.5, 61.7) |
| **Comorbidities – n (%)** |  |  |  |  |
| No comorbidity | 8 (53.3) | 8 (57.1) | 6 (54.5) | 22 (55.0) |
| At least 1 | 7 (46.7) | 6 (42.9) | 5 (45.5) | 18 (45.0) |
| **Covid-19 symptoms (most common) – n (%)** |  |  |  |  |
| Cough | 10 (66.7) | 9 (64.3) | 8 (72.7) | 27 (67.5) |
| Dyspnea | 9 (60.0) | 8 (57.1) | 9 (81.8) | 26 (65.0) |
| Fatigue | 5 (33.3) | 6 (42.9) | 3 (27.3) | 14 (35.0) |
| Loss of smell | 6 (40.0) | 3 (21.4) | 4 (36.4) | 13 (32.5) |
| Fever | 3 (20.0) | 6 (42.9) | 4 (36.4) | 13 (32.5) |
| **Onset to Randomization (days) – Median (range)** | 7.0 (1, 13) | 3.5 (1, 24) | 8.0 (1, 12) | 7.0 (1, 24) |
| **Hospitalization to Randomization (hours) – Median (range)** | 41.1 (2, 102) | 39.5 (3, 107) | 26.0 (4, 103) | 36.4 (2, 107) |
| **8-point Ordinal Scale – n (%)** |  |  |  |  |
| 4 – Hospitalized no O<sub>2</sub> | 1 (6.7) | 3 (21.4) | 1 (9.1) | 5 (12.5) |
| 5 – Hospitalized requiring O<sub>2</sub> | 5 (33.3) | 10 (71.4) | 7 (63.6) | 22 (55.0) |
| 6 – Hospitalized High flow/non-invasive | 9 (60.0) | 1 (7.1) | 3 (27.3) | 13 (32.5) |

No drug related adverse events were reported in the study. No severe (≥ Grade 3) or serious adverse events were observed in patients who received mupadolimab. Serious adverse events were reported in two patients in the placebo arm: 1 patient with acute respiratory distress syndrome and 1 patient with steroid induced psychosis.

The results for the primary and key secondary endpoints are summarized in Table 3. All endpoints trended toward more favorable outcome for mupadolimab treated patients vs. placebo. For the primary endpoint, 93.3%, 85.7% and 81.1% were alive and free from respiratory failure in the 2 mg/kg, 1 mg/kg and placebo cohorts, respectively. These differences were not statistically significant. Positive trends favoring mupadolimab also were seen for all the key secondary endpoints of time to clinical improvement, time to sustained improvement and time to discharge from the hospital. Kaplan Meier curves for these endpoints are shown in Figure 4A-4F.

**Table 3:** Summary of Efficacy Endpoints. CI – confidence interval

|  | 2 mg/kg + SOC<br>(N=15) | 1 mg/kg + SOC<br>(N=14) | Placebo + SOC<br>(N=11) |
| --- | --- | --- | --- |
| Free from Respiratory Failure or Death – (%) | 93.3 | 85.7 | 81.1 |
| Median Days to Discharge (95% CI) | 6.0 (4-12) | 4.0 (2-5) | 7.0 (2-12) |
| Median Days to Improvement (95% CI) | 7.0 (4-9) | 5.5 (3-14) | 11.0 (2-14) |
| Median Days Sustained Improve (95% CI) | 8.0 (4-12) | 6.0 (3-14) | 11.0 (2-21) |

**Fig 4.**
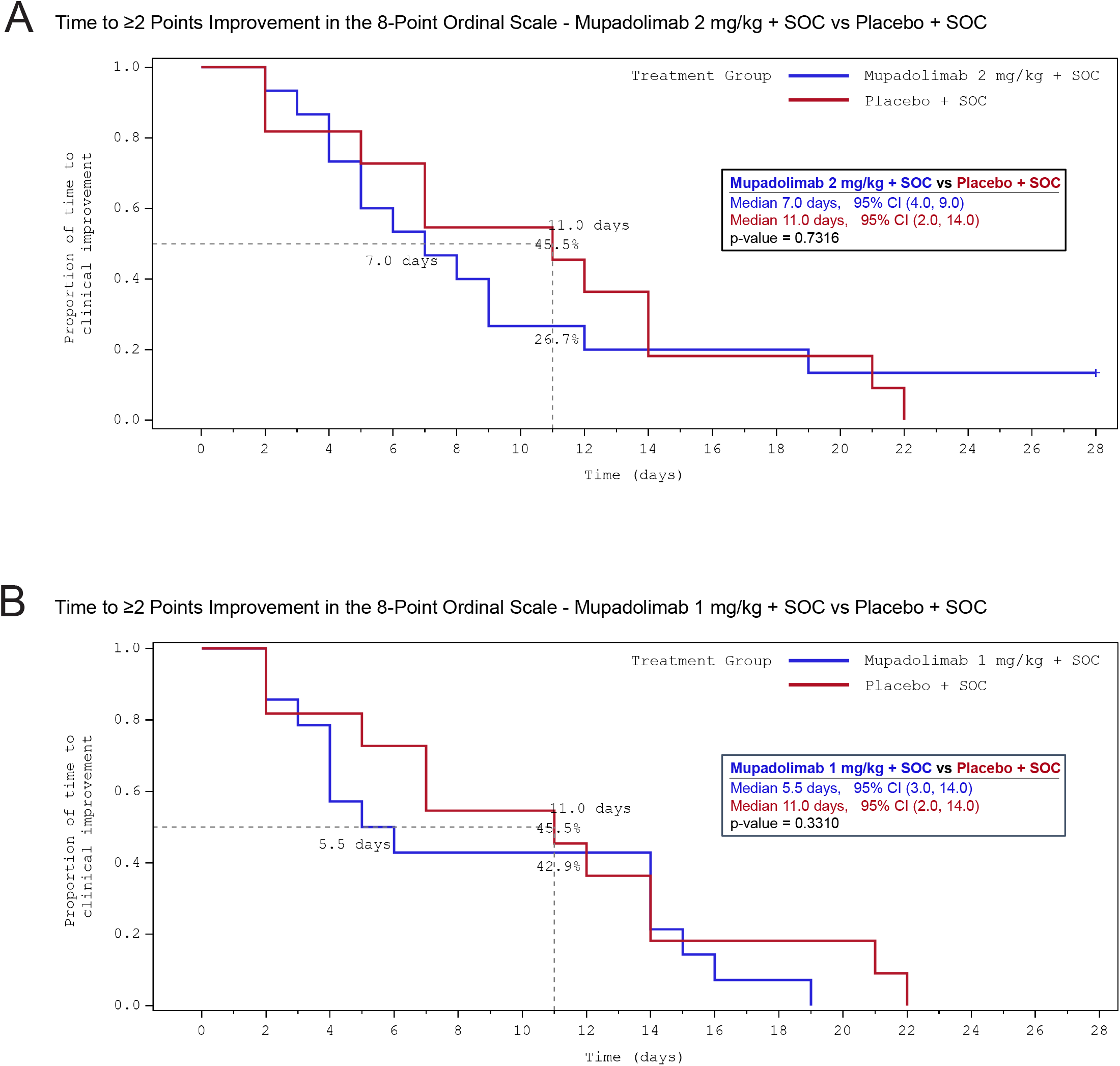
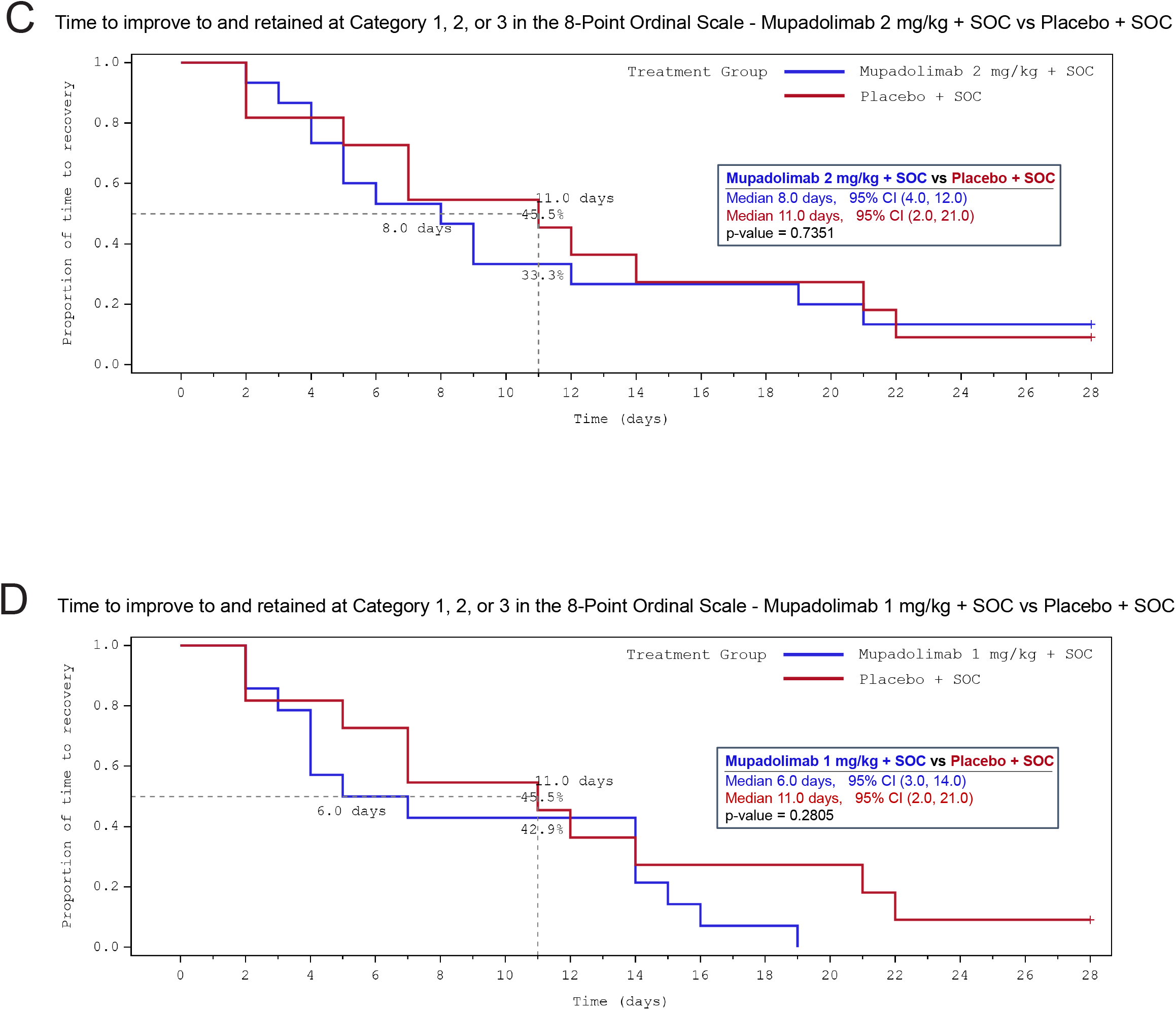
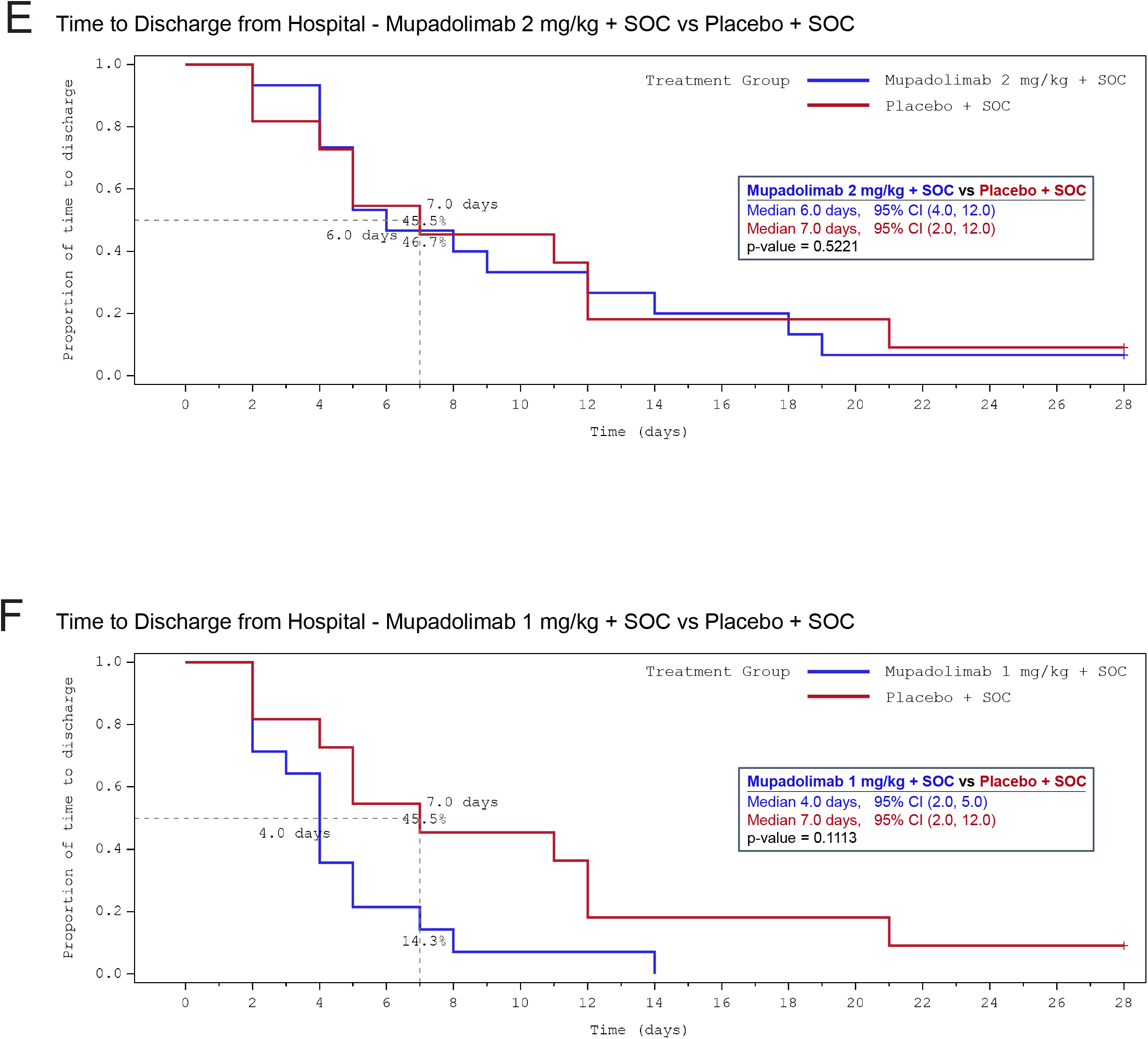
Kaplan-Meier plots of time to events for secondary endpoints compared to placebo. A-F

#### Neutralizing antibody response data

Pretreatment and day 28 serum samples were available for 26 patients: 7, placebo; 8, 1 mg/kg cohort; and 11, 2 mg/kg cohort. Antibody response to Wuhan trimeric spike protein measured by ELISA was similar between cohorts at baseline and at day 28. An assay that measures serum blocking of RBD binding to ACE2 was used to assess neutralization against Wuhan and alpha (B.1.1.7), beta (B.1.351), gamma (P.1) and delta (B.1.617.2) variants. As shown in Figure 5A, serum neutralizing titers in the 2 mg/kg cohort appeared to trend higher against Wuhan and the variants with several patients exhibiting very high titers (p=0.23 for 2 mg/kg cohort vs placebo on delta variant). Patients in the 2 mg/kg cohort also demonstrated higher cross-reactivity with the B.1.351 and P.1 variants compared to 1 mg/kg and placebo (Figure 5B). These differences were not related to duration of illness, which was similar in all groups (Table 2). We do not have the identity of the variant infecting each patient at this time.

**Fig 5.**
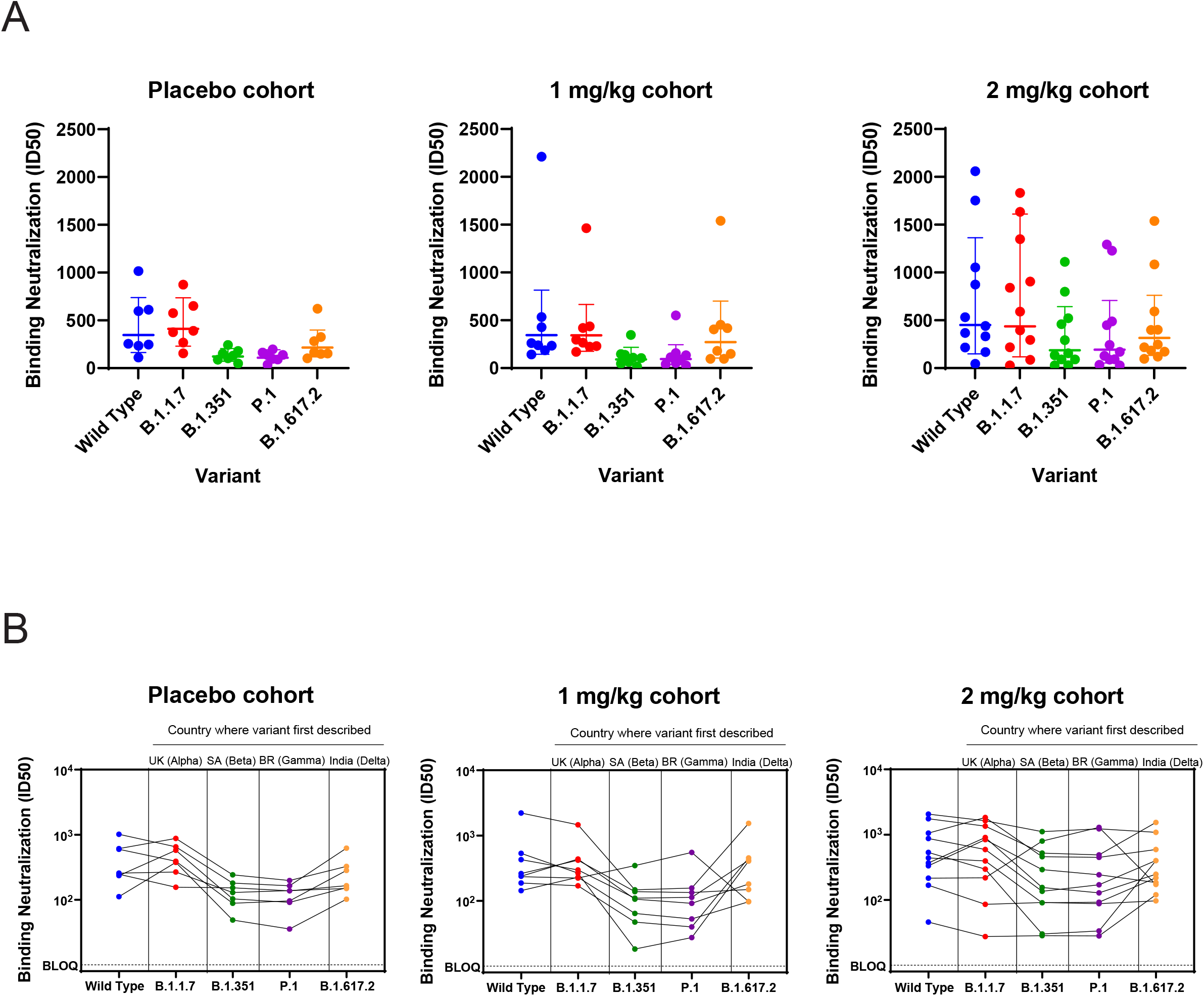
ACE2 / RBD blocking assay. A) ID50 for serum samples assayed against wild type (Wuhan) and variants using ACE2-RBD blocking assay. B) As in A, with data showing reactivity for each patient against variants.

## DISCUSSION

We have produced an anti-CD73 antibody with novel immunostimulatory properties and now report additional safety and immunologic activity in hospitalized patients with COVID-19. CD73 was originally characterized as a costimulatory molecule for T cells, but our previously reported results presented demonstrated that mupadolimab predominantly activates CD73^POS^ B cells.[17] CD73 is most highly expressed on human IgD^POS^IgM^DIM/NEG^ naïve B cells, and CD27^POS^ memory B cells expressing IgG or IgA.[11] Mupadolimab induces the expression of CD69, an activation marker that negatively regulates S1PR1 function, resulting in the prolonged retention of activated B cells in lymphoid organs and thymus.[26] This increased lymphoid residence time potentially provides time to complete B cell activation and interaction with CD4^POS^ T follicular helper cells to shape downstream immune responses. Previous studies with mupadolimab also have shown that it induces the secretion of cytokines involved in B cell differentiation including CCL22. Liu et al. have published that CCL22 promotes efficient B cell maturation in germinal centers by increasing the interaction of B cells with T follicular helper cells.[27] While B cell activation with mupadolimab is independent of adenosine blockade in vitro, the CD73 enzymatic blockade may be complementary in vivo as adenosine has been shown to restrict lymphocyte migration into lymph nodes in preclinical animal models.[28] The role of CD73 expression on cells contained in lymph nodes and in germinal centers is of potential importance.[29] Recent data shows that lymph nodes from SARS-CoV-2 infected patients lack germinal centers and would be expected to have impaired humoral immunity.[30] It is conceivable that mupadolimab restores proper immune function in these compromised germinal centers either through its action on B cells or other CD73 positive cells.

Here we report data from a completed open label phase 1 study in hospitalized patients demonstrating that antibody titers to trimeric spike protein and to the RBD of SARS-CoV-2 with neutralizing activity, increasing within 7 days in patients treated with a single dose of mupadolimab at 0.3 - 5 mg/kg. The doses evaluated in our study were selected because previous in vitro studies have shown that concentrations of mupadolimab exceeding 1 μg/ml are required for maximal B cell activation and such levels are achieved in vivo with doses of at least 1 mg/kg.

Although lacking a control arm, the median time to hospital discharge was 3 days. This compares favorably to reports from other studies in comparable patient populations.[5-8, 31] Most patients with COVID-19 become seropositive for IgG/IgM/IgA within 2-3 weeks following onset of symptoms.[32] The patients in the Phase 1 trial had low levels of anti-SARS-CoV2 antibodies at the time of hospitalization despite a wide range of duration of prior symptoms from 1-21 days. The lack of response in these patients may be related to host factors reducing the ability to mount a humoral response or other unknown immunosuppressive effects of viral infection. Mupadolimab may overcome this apparent immunodeficiency as we observed robust anti-SARS-CoV-2 antibody responses induced by mupadolimab in patients with long period POS and low pre-treatment titers. The kinetics following seroconversion are still being clarified, but others have reported that antibody titers in COVID-19 patients plateau approximately 6 days after seroconversion before steadily declining in the following weeks (IgM/IgA) to months (IgG).[32] Our results show that anti-SARS-CoV2 titers continue to rise reaching a stable plateau in the weeks following treatment, possibly hinting at a more robust and durable humoral response that would theoretically improve clinical outcomes in COVID-19 patients. These effects may also serve to reduce viral transmission. In line with this idea, Chen et al. have shown that sustained antibody titers predict favorable clinical outcome in COVID-19.[33]

Based on these findings, a randomized double-blind, placebo-controlled trial was undertaken, but was discontinued early, after enrolling 40 patients, due to waning of the COVID-19 incidence in July 2021. Single injections of either 2 mg/kg or 1 mg/kg had no reported adverse events, with two serious adverse events seen in the placebo group. These adverse events consisted of acute respiratory distress syndrome and steroid psychosis, both reported as disease related. Although stopped prematurely and therefore underpowered to show statistical significance, the primary and secondary endpoints all trended towards favoring the treatment cohorts. The largest impact on the primary endpoint was seen in the 2 mg/kg cohort despite this group having patients with more advanced disease on enrollment. In a phase 3 study in hospitalized patients using convalescent plasma, the median time to discharge was 13 days.[31] In a recent Phase 3 trial in hospitalized patients with COVID-19 treated with tofacitinib, the proportion of patients progressing to respiratory failure or death was 18% for tofacitinib and 29% in the placebo group. Further studies will be required to establish the safety and efficacy of mupadolimab. We believe the results from our studies will encourage additional clinical testing in COVID-19. The effects seen with relatively low doses of mupadolimab indicate that alternative routes of delivery such as subcutaneous or intramuscular administration are feasible. Such low doses and routes of administration may make this treatment feasible in the outpatient setting or possibly delivered as an adjuvant for vaccination of healthy subjects.

We evaluated antibody responses to SARS-CoV-2 spike protein in our treated patients. Serum binding to Wuhan trimeric spike protein reached high titers in ELISA in all our treated patients. In the Phase 3 trial, we tested serum for neutralization activity using a blocking assay that has been shown to strongly correlate with viral neutralization. Patients in this trial were enrolled prior to the emergence of the delta variant in the U.S. Patients receiving 2 mg/kg of mupadolimab demonstrated a trend toward higher titers against Wuhan and each of the variants. These results are consistent with the proposed mechanism of B cell activation and induction of broadly cross-reactive antibodies. Such responses could be protective against future infection with other variants. In addition to a potential role in stimulating humoral immunity, B cells are known to be important in antigen presentation and induction of T cell immunity.[34]

Multiple viral and mRNA-based vaccines in development have demonstrated the potential to induce humoral and cellular responses to SARS-CoV-2 antigens in vaccinated normal subjects.[35, 36] While encouraging, diminishing antibody titers, especially to variants, have necessitated booster vaccinations.[37, 38] Mupadolimab is being developed as a therapeutic, but it is important to note that it could potentially be used as a vaccine adjuvant to boost the titer, diversity, and duration of antibody responses and potentiate the development of long term immunity by generating memory B and effector T cells. This combination approach may be useful to enable successful immunization with a single vaccination and particularly effective in vulnerable populations such as immunosuppressed and elderly patients who do not typically respond well to vaccines.

Absent a larger randomized trial, we are currently unable to conclude that the effects observed thus far in treated COVID-19 patients are directly attributable to mupadolimab. Our current results demonstrate an atypically robust and durable antibody response following mupadolimab treatment, but we cannot exclude the possibility that these responses would have developed naturally. Moreover, multiple confounding factors including the variability in patient responses, clinical setting, and lack of standardized testing methods complicate comparisons to other therapeutics in development. Nonetheless, these encouraging early results are in line with our hypothesized biological mechanism. If confirmed, mupadolimab could be useful for the treatment or prevention of other viral infections and future pandemics.

## METHODS & METHODS

### Preclinical mouse immunization model

NSG-SGM3 mice (Jackson Laboratories, stock #013062) were immunized with an emulsion of 50 ug full length spike protein from SARS-CoV-2 (ABClonal) and Freund’s Incomplete Adjuvant (Sigma) subcutaneously on both the left and right flank (25 μg/side). Mice were then randomized into two treatment groups: half the animals were dosed daily (i.p.) with 10 mg/kg mupadolimab, and half the animals were dosed daily with 10 mg/kg hIgG1 (BioXCell). Animals were cheek bled on days 1 (pre-bleed), 4, 8, 11 and 15 to assess anti-spike or anti-nucleocapsid (as a negative control) antibody production over time by ELISA.

### Anti-SARS-CoV-2 antibody ELISA assays

ELISA was performed to measure the IgG and IgM to the receptor-binding domain (RBD) of the spike protein, full-length spike trimer of the SARS-CoV-2 virus. Purified recombinant SARS-CoV-2 RBD and full-length spike protein were obtained from Lake Pharma. ELISA plates were coated with RBD or spike protein (2 μg/mL in PBS) overnight at 4°C then blocked with 3% BSA in PBS for 1 hr. at room temperature (RT) after three washes with PBST. Serial dilutions of serum were prepared in PBST containing 1% BSA, then dispensed to the wells of the coated microtiter plate and incubated for 2 hr at RT. After three washes, the bound antibody was detected using anti-human IgG-horseradish peroxidase (HRP) conjugated secondary antibody (1:3000, Sigma-Aldrich, A0170) or anti-human IgM HPR secondary antibody (1:3000, Sigma-Aldrich, A0420). After three washes, the reaction was developed by the addition of the substrate o-phenylenediamine dihydrochloride (SigmaFast OPD) and stopped by HCl (2M). The absorbance at 490 nm (OD490) was measured using an Envision plate reader (PerkinElmer). Recovered COVID-19 patient serum was obtained from Sanguine Biosciences from recovered patients confirmed to have had a COVID-19 PCR+ result. All serum samples were treated to inactivate infectious virus by incubation at 56°C for 30 mins. Healthy volunteer serum samples obtained from Stanford Blood Center during 2018 served as negative control. The titer cutoff value at OD490 is the mean plus 3 standard deviations of the negative controls. The endpoint titer is reported as the highest dilution of at least two before the OD490 decreases below the cut-off value. Data were analyzed using GraphPad Prism 7.

### Neutralizing Antibody ELISA

Neutralizing antibody titers in a biochemical ELISA format were measured using the MSD platform. Briefly, human sera were prepared using a 3-fold serial dilution starting at 1:30. The diluted sera (25 µL) was transferred to the wells of a blocked multiplex plate precoated with wild type and variant spike proteins (MSD) and incubated for 60 min at room temperature. The sera was diluted a further 2-fold by addition of an equal volume of sulfotagged ACE2 (MSD) and the mixture incubated for 60 min at room temperature. Unbound sulfotagged ACE2 was removed by washing the plate three times with 150 µL of phosphate-buffered saline, 0.05% Tween-20. Bound ACE2 was detected by addition of 150 µL of 1X read buffer (MSD) and chemiluminescence read using a Quickplex SQ 120 plate reader (MSD). ID50 values were obtained by fitting the response data to a four-parameter logistic equation using GraphPad Prism version 8.4.3 for Windows, GraphPad Software, San Diego, California USA.

### Viral neutralization assays

SARS-CoV-2 spike pseudotyped lentivirus were produced with the Wuhan-Hu-1 (wild type), B.1.1.7 or B.1.351 variant spike as the envelope glycoprotein and the firefly luciferase gene as a reporter (BPS Bioscience). Neutralization activity was determined using heat inactivated serum (56°C, 30 min) mixed with pseudovirus before addition to HEK293T-hACE2 cells. Luciferase activity was measured seventy-two hours after transduction and SARS-CoV-2 neutralization titers were defined as the reciprocal dilution yielding a 50% reduction in signal relative to the average of uninhibited control wells.

## Data Availability

Further information and requests for resources and/or reagents should be directed to Dr. Richard A. Miller.

## ACKNOWLEDGEMMENTS

The authors wish to thank hospital staff caring for these patients, and the patients and their families for participating in these studies.

